# Reverse Inflammaging: Long-term effects of HCV cure on biological age

**DOI:** 10.1101/2022.03.02.22271777

**Authors:** Carlos Oltmanns, Zhaoli Liu, Jasmin Mischke, Jan Tauwaldt, Yonatan Ayalew Mekonnen, Melanie Urbanek-Quaing, Jennifer Debarry, Benjamin Maasoumy, Heiner Wedemeyer, Anke R.M. Kraft, Cheng-Jian Xu, Markus Cornberg

## Abstract

**Background and Aims:** Chronic hepatitis C virus (HCV) infection can be cured with direct-acting antiviral agents (DAA). However, not all sequelae of chronic hepatitis C appear to be completely reversible after sustained virologic response (SVR). Recently, chronic viral infections have been shown to be associated with biological age acceleration defined by the epigenetic clock. The aim of this study was to investigate whether chronic HCV infection is associated with epigenetic changes and biological age acceleration and whether this is reversible after SVR.

**Methods:** We included 54 well-characterized patients with chronic hepatitis C at three time points: DAA treatment initiation, end of treatment, and long-term follow-up (median 96 weeks after end of treatment). Genome-wide DNA methylation status from peripheral blood mononuclear cells (PBMC) was generated and used to calculate epigenetic age acceleration (EAA) using Horvath’s clock.

**Results:** HCV patients had an overall significant EAA of 3.12 years at baseline compared with -2.61 years in the age-matched reference group (p<0.00003). HCV elimination resulted in a significant long-term increase in DNA methylation dominated by hypermethylated CpGs in all patient groups. Accordingly, EAA decreased to 1.37 years at long-term follow-up. The decrease in EAA was significant only between the end of treatment and follow-up (p=0.01). Interestingly, eight patients who developed hepatocellular carcinoma after SVR had the highest EAA and showed no evidence of reversal after SVR.

**Conclusions:** Our data contribute to the understanding of the biological impact of HCV elimination after DAA and demonstrate that HCV elimination can lead to “reverse inflammaging”. In addition, we provide new conceptual ideas for the use of biological age as a potential biomarker for HCV sequelae after SVR.

**Lay Summary:** Chronic hepatitis C virus infection is now curable with direct acting antiviral agents (DAA), but are concomitant and sequelae also fully reversible after cure? Recent data demonstrate that chronic viral infections lead to an increase in biological age as measured by epigenetic DNA methylation status. Using a unique cohort of hepatitis C patients with and without cirrhosis as well as progression to HCC, we demonstrated that these epigenetic changes and concomitant increase in biological age are also observed in chronic HCV infection. Our data further suggest that this effect seems to be partially reversible in the long-term course after sustained virological response (SVR) by DAA therapy and that biological regeneration occurs. In this regard, the recovery effect appears to be dependent on disease course and was significantly lower in patients with progression to HCC. This suggests the use of biological age based on epigenetic state as a potential biomarker for HCV sequelae.

**Graphical abstract:** 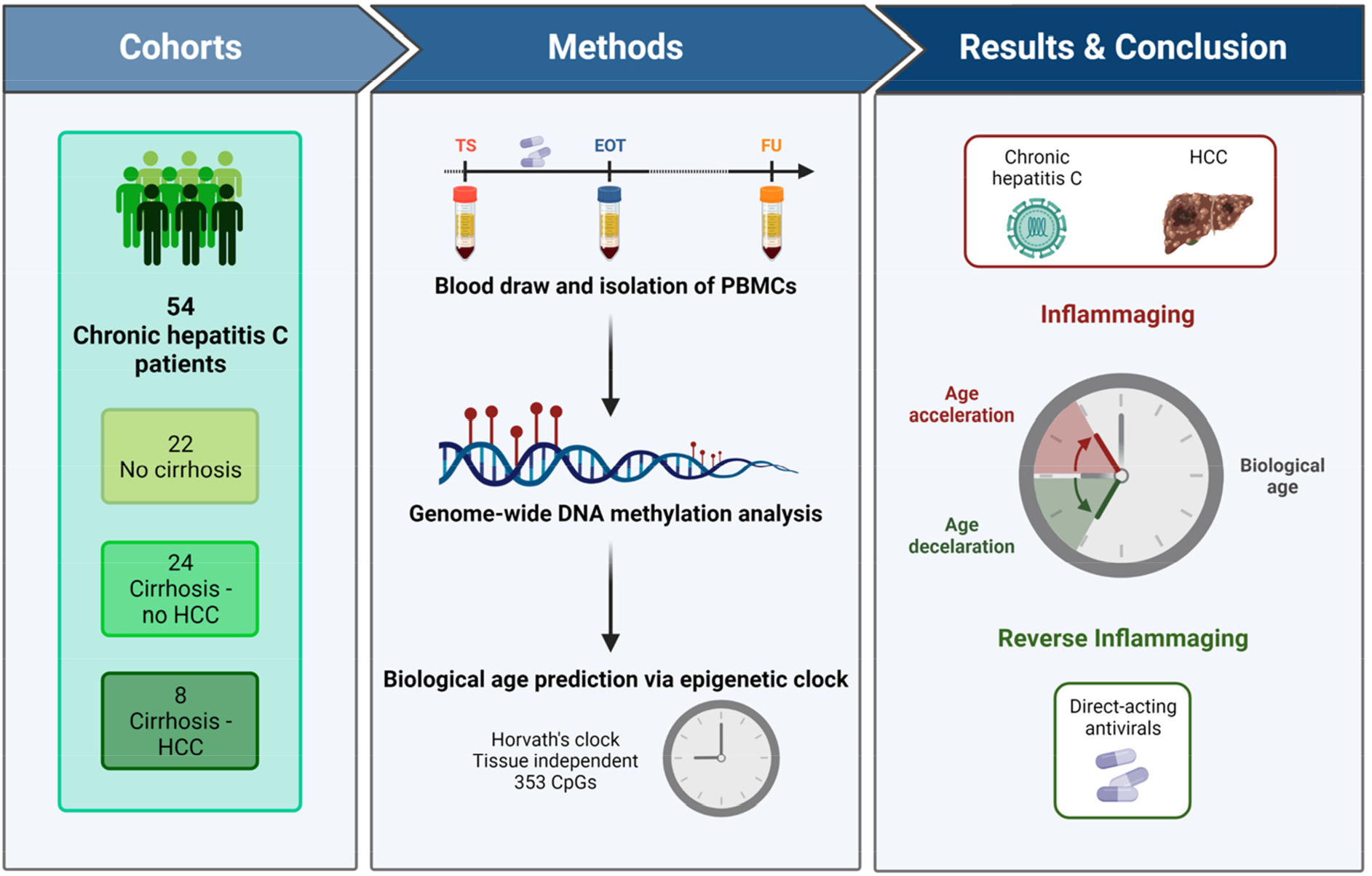

**Highlights:**

- Patients with chronic hepatitis C have accelerated epigenetic age compared with healthy controls.
- DAA treatment and HCV elimination partially reverse the accelerated epigenetic age in the long-term follow-up.
- Patients who developed hepatocellular carcinoma after HCV elimination did not show reversal of accelerated epigenetic aging during the follow-up.

## Introduction

Hepatitis C virus (HCV) infection continues to be a major global health burden. According to the World Health Organization (WHO), 1.5 million new HCV infections occur worldwide each year, with 58 million individuals living with chronic HCV infection [1]. The main long-term consequence of chronic hepatitis C is the development of liver cirrhosis, which is associated with a significant risk of hepatocellular carcinoma (HCC) [2]. Furthermore, HCV infection can lead to extrahepatic manifestations such as chronic fatigue, diabetes mellitus or vasculitis [3]. As a result, approximately 290,000 people die each year as a result of HCV infection [1].

Meanwhile, direct acting antivirals (DAA) are available, well tolerated and result in sustained virological response (SVR) rates of more than 95%, leading to a significant reduction of liver morbidity and mortality [4]. However, not all sequelae of chronic hepatitis C seem to be completely reversible after SVR. Patients with advanced fibrosis or cirrhosis have a residual risk for HCC [5]. Impaired quality of life [6] or extrahepatic manifestations such as cryoglobulinemic vasculitis [7] are only partially improved or not reversible in all patients. Interestingly, recently published studies have shown that HCV infection can leave an immunological imprint or scar after SVR, and the impaired immune response characteristic of chronic HCV infection is only partially restored [8] [9] [10].

One mechanism that may explain this is that HCV infection can profoundly affect the epigenome, and it has been shown that much of the epigenetic changes caused by HCV remain as “scars” in different cell types, i.e. CD8 T cells and hepatocytes, after viral elimination [11][12]. Of note, HCV induced epigenetic changes in hepatocytes have been associated with HCC risk and this persisted after SVR [13]. DNA methylation (DNAm), defined as the covalent addition of a methyl group to a DNA nucleotide (usually the cytosine of a cytosine-guanine dinucleotide [CpG]), is the most well-studied epigenetic modification that affects transcription factor binding and controls accessibility to regulatory regions in the DNA, modulating gene expression [14].

Numerous studies have shown that epigenetic changes, particularly DNAm, are affected by aging [15] [16], so DNAm status enables us to estimate an individual’s biological age. One of the most widely used and validated methods for estimating biological age is “Horvath’s clock,” which is based on the methylation status of 193 CpGs that gain methylation and 160 CpGs that lose methylation over time [17].

Thus, the aim of this study was to evaluate if chronic HCV infection is associated with biological age acceleration and if this is reversible after DAA therapy and HCV elimination. For this, we analyzed DNAm on peripheral blood cells isolated from a well-characterized cohort of 54 patients with chronic hepatitis C before, at the end and in the long-term follow-up after DAA therapy.

## Methods

### Study population and design

Out of 799 patients with chronic HCV infection treated with DAA at Hannover Medical School between January 2014 and March 2021, a total of 54 well-characterized patients were selected for this study (Supplementary Figure 1). The patients are part of a biobank registry and all patients gave written informed consent. Blood samples were collected and Blood samples were collected and stored according to established Standard Operation Procedures (SOP).

The study protocol conformed to the ethical guidelines of the Declaration of Helsinki and the local ethics committee approved this study a priori (Nr. 9474_BO_K_2020).

The study comprises 3 different groups of patients. Cohort A contains 22 patients without liver cirrhosis. Exclusion criteria for this group are depicted in Supplementary Figure 1. The sampling time points for this group included the start of therapy, the end of treatment, and a follow-up period of 96 weeks. Cohort C includes 8 patients who developed HCC after SVR. These 8 patients were matched with 24 other patients (cohort B) using a propensity score (PS) approach [18]. So the total cohort includes 32 patients with cirrhosis. Sampling time points included initiation of therapy, end of treatment, and last available sampling time point (before the development of HCC in the HCC group). The detailed baseline characteristics of the patients are shown in Table 1.

**Table 1:**
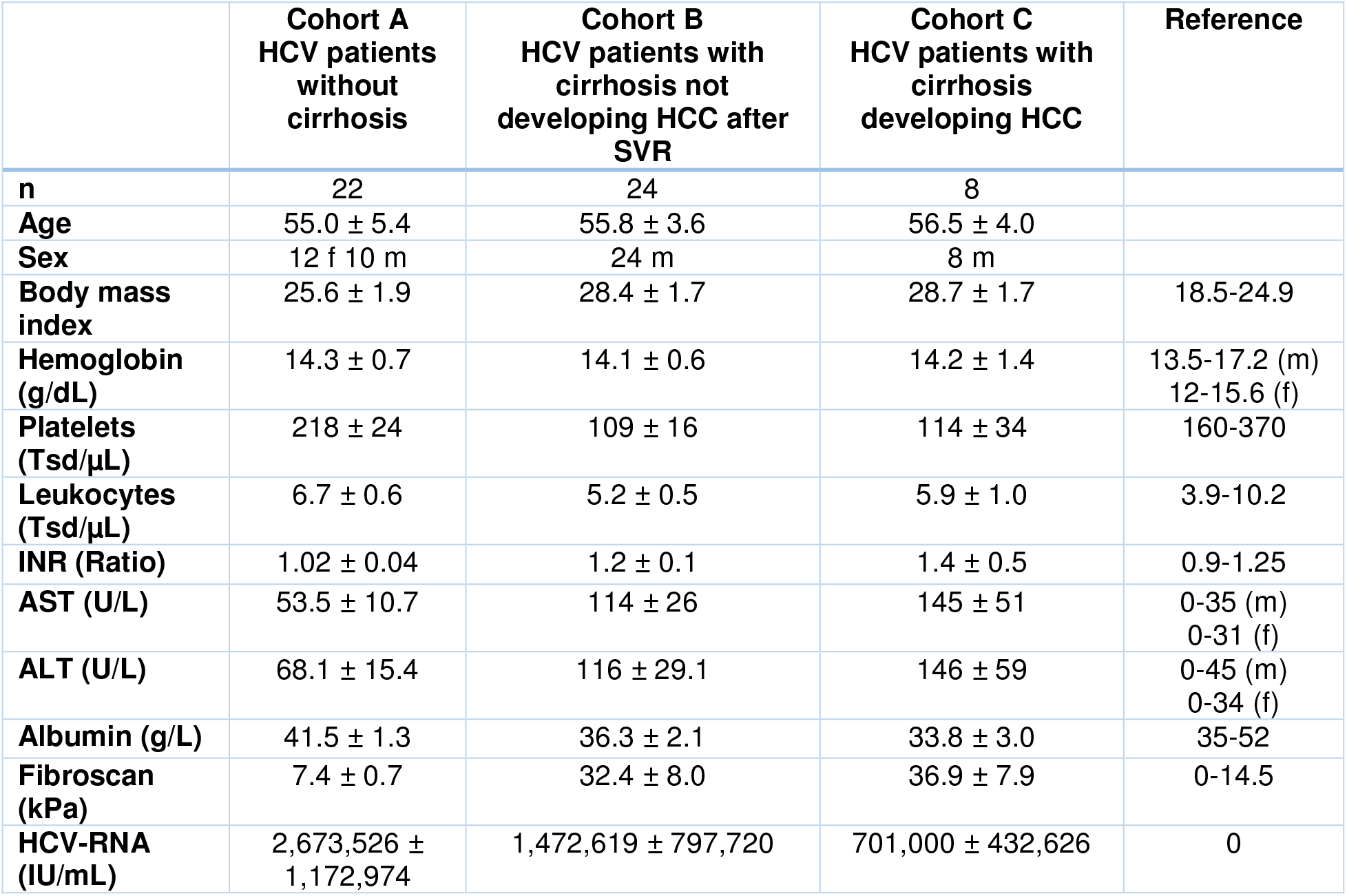
Baseline characteristics of analyzed HCV cohorts. Clinical characteristics of different subgroups including: HCV patients without cirrhosis (cohort A), HCV patients with cirrhosis not developing HCC after SVR (cohort B) and HCV patients with cirrhosis developing HCC (cohort C).

**Table 2:**
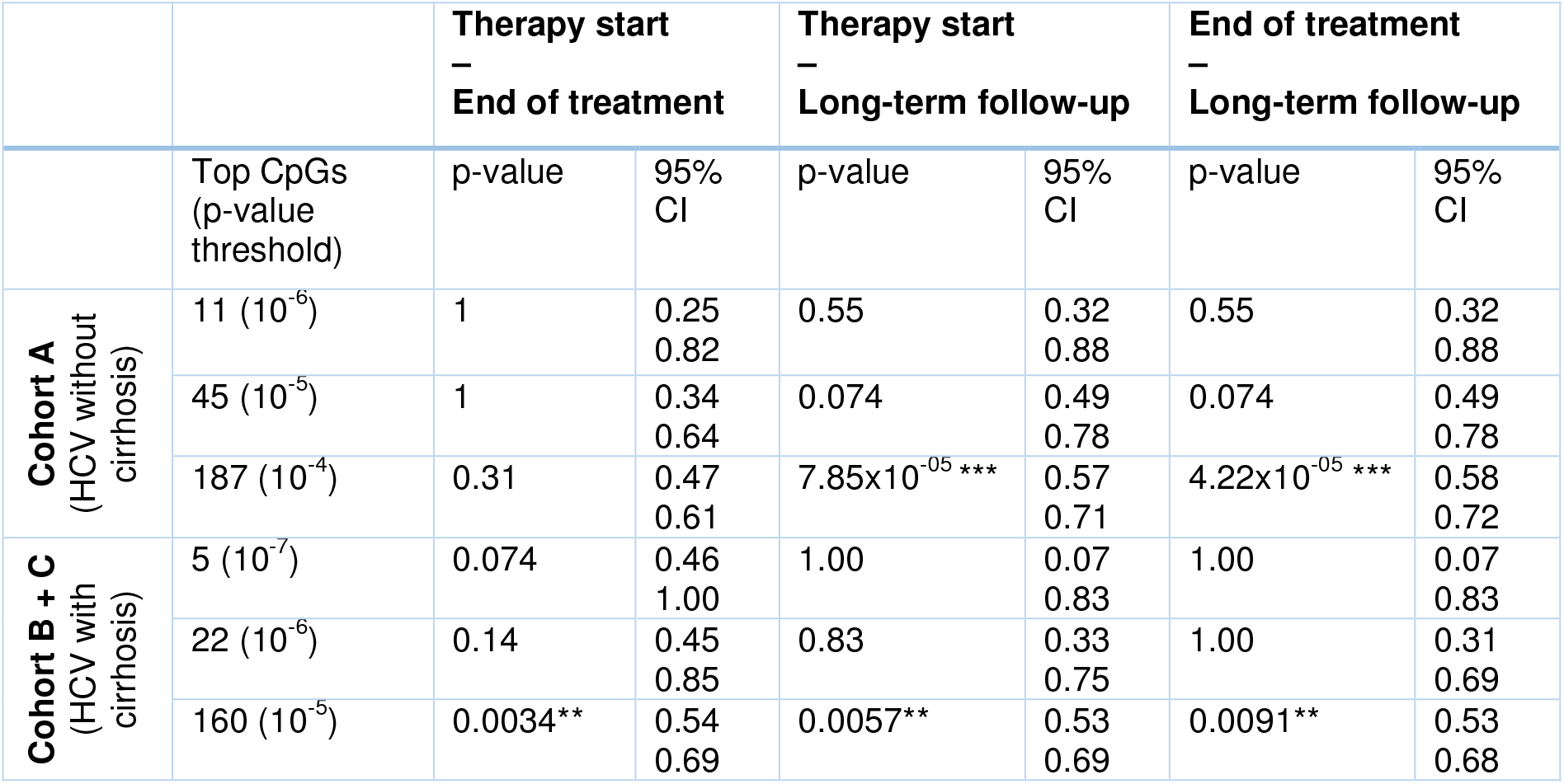
Global methylation trends in HCV cirrhosis (cohorts B and C) and non-cirrhosis (cohort A) patients. Two-sided proportion test was used to determine whether in- and decrease in methylation differed at top p-value thresholds.

### Sample preparation

Peripheral blood was drawn from patients at the Hannover Medical School outpatient clinic at the indicated time points, and peripheral blood mononuclear cells (PBMCs) were isolated according to a standard Ficoll Hypaque density centrifugation protocol (BioColl separating solution; Biochrom AG, Berlin, Germany). After isolation, cells were transferred to a freezing medium and stored in liquid nitrogen. For analysis purposes, the cells were thawed, counted and then further processed by us using the Monarch Genomic DNA Purification Kit T3010L (New England Biolabs, Ipswich, MA, USA) according to manufacturers instructions. We normalized the DNA concentration of all samples after elution to 60 ng/µL, randomized the samples on a 96-well plate and stored the plate at -20 degrees Celsius.

### Cohort Matching

Cohorts B and C were matched using R Statistical Software (Version 4.0.5.) and the MatchIt package [18]. Nearest neighbor matching was used as the method of matching [19]. Matching variables included sex, age, transient elastography and follow-up length. Following the matching process, we checked for significant differences between the HCC and the control group. There were no significant differences detectable in the matching variables.

### DNA methylation measurements and quality control

DNA methylation measurement was performed at the Human Genomics Facility of Erasmus MC, Rotterdam, the Netherlands. 500ng DNA was bisulfite converted using the EZ-96 DNA Methylation kit (Zymo Research Corp., Irvine, CA, USA) with the KingFisher Flex robot (Thermo Fisher Scientific, Breda, the Netherlands). Methylation was assessed for 54 individuals in 8 µL bisulfite treated DNA using the Infinium MethylationEPIC BeadChip, following Illumina’s protocol.

After receiving the raw IDAT files, we started our data preprocessing for each cohort in “R Statistical Software” using the Bioconductor package minfi [20]. We performed an extensive quality control by checking for gender concordance and removing low-quality probes (detection p-value > 0.01 in more than 10% of all samples), SNP-containing probes, cross-reactive probes [21] and sex chromosomal probes. All samples were checked for 20 control metrics generated by the “BeadArray Controls Reporter Software” [22]. From the 162 samples, we excluded three samples not passing the pre-defined cut-off values by Illumina indicating a failed bisulfite conversion.

In the first and second batch of samples, we excluded 1,470 and respectively 1,302 failed probes, 43,254 cross-reactive probes [21], 19,627 sex chromosome probes and 11,681 probes with SNPs at the CpG interrogation or at the single nucleotide extension. Overall, we included 793,532 and 793,365 unique high-quality CpGs from two batches respectively and 162 samples in our analysis.

For normalization of the data, we used “dasen” from the “wateRmelon” R package [23]. We used the “IlluminaHumanMethylationEPICanno.ilm10b4.hg19” package to annotate all CpGs passing our quality control. In our downstream analysis we used M values for all of our analysis.

### Biological age prediction

We used “agep” function from “wateRmelon” R package [23] to predict the biological (epigenetic) age of samples using Horvath’s coefficients. Epigenetic age acceleration (EAA) was defined as the difference between epigenetic and chronological age. The age-matched healthy cohort from a publicly available dataset (GSE40279) [24] has been used to obtain the expected distribution of EAA in healthy population. Cell proportion was obtained via https://dnamage.genetics.ucla.edu/new.

### Global methylation trends

We aimed to understand the global methylation trends by taking a look at the most changed CpGs over course of treatment and follow-up. To detect those differences we designed a linear mixed effects model (lmer(methylation ∼ time point + AST + CD8 T-cells + CD4 T-cells + NK cells + B cells + (1|Patient ID)), which included inflammation as well as immune phenotype parameters as covariates. Finally, two-sided proportion test was used to determine whether in- and decrease in methylation differed at fifferent p-value thresholds.

### Statistical analysis

We used R Statistical Software (Version 4.0.5.) to conduct our analyses. Based on this, we created all graphics in either R with “ggplot” and “ggpubr” packages or in GraphPad Prism (Version 8.3.1.). For detection of differences between dependent sample groups, “Wilcoxon signed-rank test” was used. Correspondingly, we used “Mann-Whitney U test” for differences between independent sample groups. “Spearman” method was used for calculating the correlation between EAA and estimated cell proportions as well as clinical lab parameters. P-values. P-values for correlations between EAA and clinical lab parameters were adjusted using false discovery rate.

## Results

### Characteristics of the study cohort

Patients in cohort A (no cirrhosis) included 12 females and 10 males with a mean age of 55.0 years. AST and ALT levels at baseline were elevated and declined during treatment and all patients had normal levels at follow-up (Supplementary Figure 2). All patients of this cohort had a transient elastography (FibroScan) value of less than 11.0 kPa indicating that no liver cirrhosis was apparent at treatment start. Patients in cohorts B and C (HCC after SVR and matched controls without HCC) were all male and had significantly lower platelet counts, albumin and INR levels (Table 1). In addition, the patients had higher levels of AST, ALT and higher transient elastography (FibroScan) values, which also declined during therapy (Supplementary Figure 2). At follow-up, 26/32 patients in cohorts B and C had normal ALT level.

Importantly, the propensity score matched control cohort B (no HCC) showed no significant differences in all lab parameters compared to cohort C (HCC after SVR), allowing a solid comparison. All patients were HCV-RNA positive at treatment start but reached SVR.

### Patients with chronic hepatitis C show an accelerated epigenetic age

To understand the impact of chronic HCV infection on biological aging, we calculated epigenetic age of all HCV patients and their age-matched control by using Horvath’s epigenetic clock. Horvath’s clock showed strong correlations between biological and chronological age, independent of the analyzed time point (p<2.2×10^−16^) (Figure 1).

**Figure 1:**
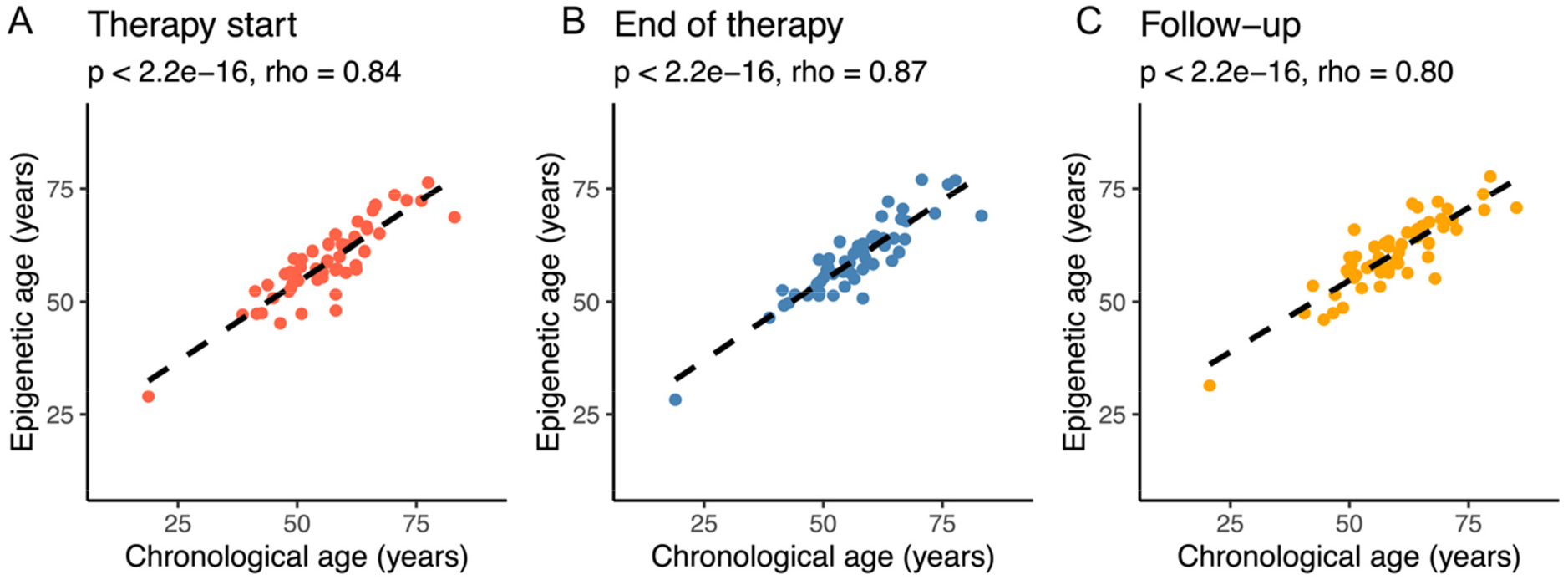
Scatter plots showing the correlation between epigenetic age (Horvath’s clock) and chronological age in chronic HCV patients at therapy start (A), end of therapy (B) and long-term follow-up (C). The rank-based Spearman method was used for calculating the correlation.

HCV patients showed an accelerated epigenetic age (EAA) at treatment start (median EAA = 3.12 years) compared to our age-matched reference group (median EAA = -2.61 years) (p=2.96×10^−5^) (Figure 2A). Further analysis revealed that EAA differed among the three patient groups studied. While chronic HCV patients without cirrhosis tended to have the lowest age acceleration at baseline (p=0.004, median EAA = 2.45 years), patients who developed HCC after SVR tended to have the highest age accelerations (p=0.004, median EAA = 4.85 years) (Figure 2B).

**Figure 2:**
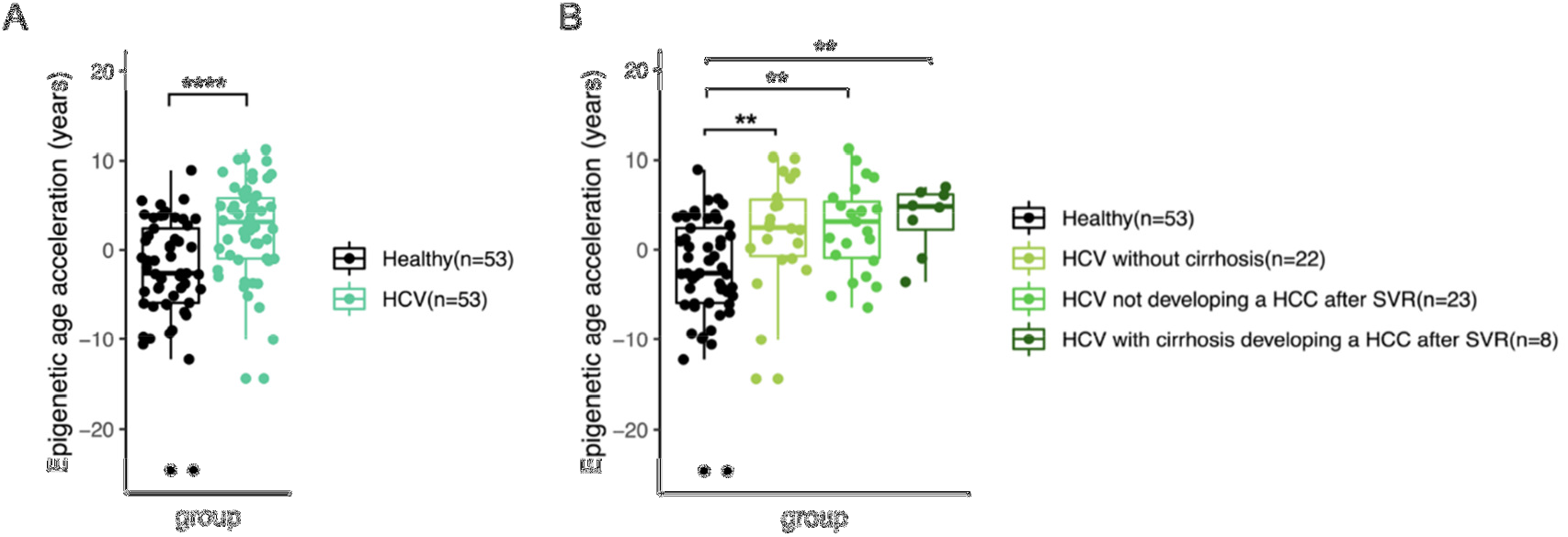
Epigenetic age acceleration compared to the reference group and across different subgroups in chronic HCV patients at therapy start. Comparison between healthy controls and HCV patients overall (A) as well as HCV patients without cirrhosis (cohort A), HCV patients with cirrhosis not developing HCC after SVR (cohort B) and HCV patients with cirrhosis developing HCC (cohort C) (B). Wilcoxon rank sum test was used to calculate the difference between groups. *: p < 0.05, **: p < 0.005, ***: p < 0.0005, ****: p < 0.00005.

### Epigenetic age acceleration decreases after DAA therapy

Our longitudinal analysis over the course of DAA treatment and long-term follow-up showed that EAA is decreasing from baseline until the long-term follow-up (Figure 3A). While the median age acceleration at baseline was 3.12 years, it decreased to only 1.37 years at long-term follow-up (n=54, p=0.07). The decrease in EAA was particularly significant between the end of therapy and the long-term follow-up (end of treatment – long term follow-up: p=0.01), whereas there was no significant difference in EAA between baseline and end of treatment (p=0.56) (Figure 3).

**Figure 3:**
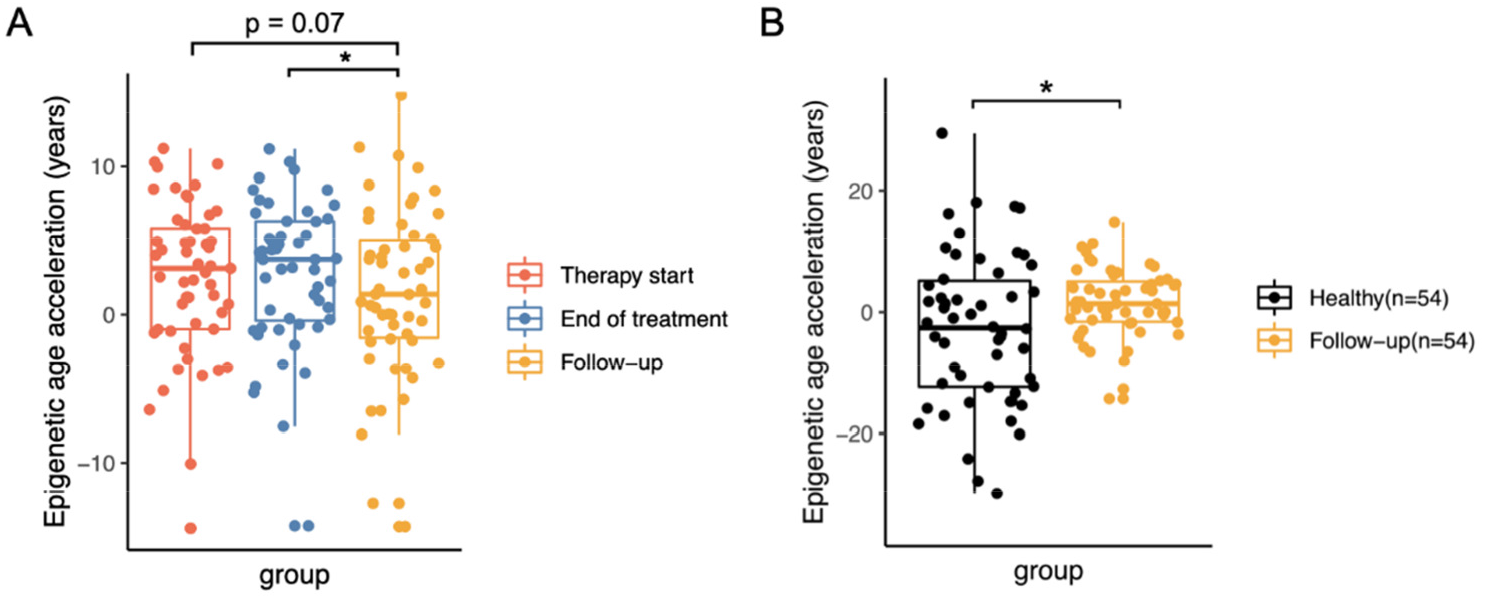
Boxplots showing the age acceleration in chronic HCV patients (n=54) at different time points over course of treatment and follow-up and age-matched healthy controls. Comparison between different sampling points (A) and HCV patients at long-term follow-up and age-matched healthy controls (B). Two-sided Wilcoxon signed-rank test was used to calculated the difference between different sampling points and two-sided Mann-Whitney U test was used for comparison of follow-up with healthy controls. *: p < 0.05.

Interestingly, the patients who developed HCC after SVR showed not only the highest EAA but also did not show a significant decline of EAA after HCV elimination (therapy start – long-term follow-up: p=0.51, median at therapy start: 4.85 years, median at long-term follow-up: 3.76 years). In contrast, PS matched patients without HCC after SVR showed a significant decline of EAA from baseline to the last follow-up (Figure 4).

**Figure 4:**
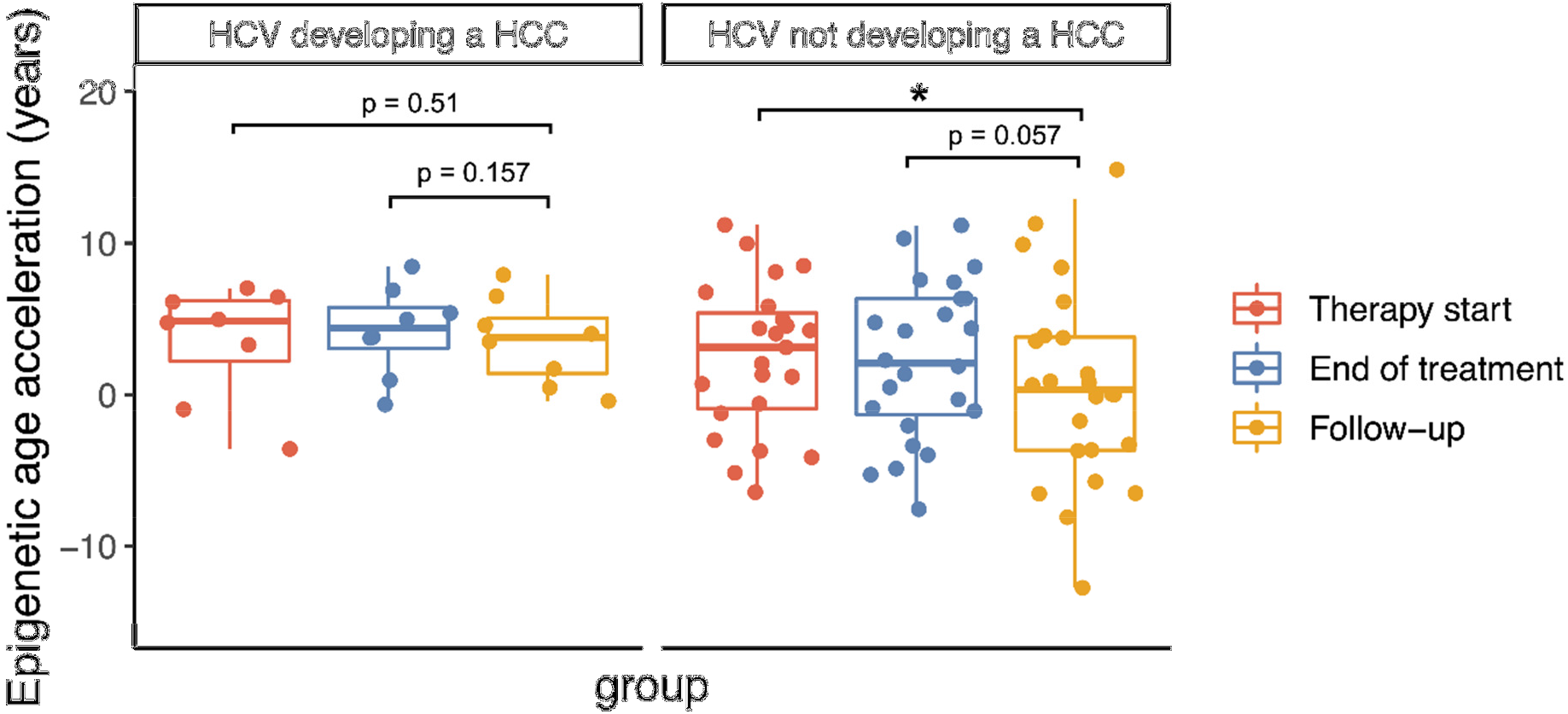
Boxplots of EAA in patients developing a hepatocelluar carcinoma after SVR and matched control group over course of therapy. One-sided paired t-test was used to calculate the difference between groups since EAA was normally distributed. *: p < 0.05

### Significant increase in DNA methylation, dominated by hypermethylated CpGs, after DAA therapy

To understand what drives the DNA methylation aging changes, we next investigated the effect of DAA therapy or HCV elimination on global methylation levels. It has been reported that overall DNA methylation levels are related to aging and in principle decrease with increasing age [25]. Interestingly, we observed that HCV elimination by DAA therapy leads to an increase in DNA methylation, which is inconsistent with the previous findings that methylation is inversed correlated with aging. This may suggest that DAA therapy has effects on epigenetic aging.

Specifically, in cohort A consisting of chronic HCV patients without cirrhosis, the treatment with DAA did not result in an overall increase in DNA methylation between therapy start and end of treatment. In contrast, there was a significant increase in methylation dominated by hypermethylated CpGs between therapy start and follow-up 96 weeks. This increase was consistent at different chosen p-value thresholds (p-value threshold 10^−4^: p=7.85×10^−5^). In the chronic HCV patients with cirrhosis (cohorts B and C) there was a significant increase in methylation already between therapy start and end of treatment (p-value threshold 10^−5^: p=0.003) that stayed consistent between therapy start and long-term follow-up (p-value threshold 10^−5^: p=0.006). Similarly compared to cohort A, this increase in methylation was mainly based on hypermethylated CpGs.

### Epigenetic age acceleration is not associated with estimated cell proportion but clinical phenotypes

As epigenetic changes can be driven by changes in cell composition, we next associated estimated cell counts with EAA in chronic HCV patients. In our approach, we focused on main cell types with high overall frequencies. In conclusion, we were not able to detect any significant correlations between EAA and estimated cell counts before, during and after treatment.

The strongest association is between monocytes and EAA at follow-up. There is a consistent trend between CD8 T cells and EAA at all sampling points which is not significant after all. No consistent pattern was detectable for all other cell types. This stayed consistent in all subgroups of our analyses (Supplementary Figure 3-5).

We also aimed to understand if epigenetic age is driven by any specific clinical phenotype. We focused on clinical lab parameters that describe liver function such as ALT, albumin, platelets, transient elastography (FibroScan) values and GGT, as well as leukocytes and creatinine. Liver stiffness, as indicated by transient elastography (FibroScan), was positively correlated with epigenetic age acceleration (adj. p=0.03, rho=0.24) in HCV patients with cirrhosis (cohorts B+C), while platelet counts were negatively correlated (adj. p=0.03, rho=-0.27) in the same cohorts. In the HCV patients without cirrhosis (cohort A) we did not observe any significant correlations (Figure 6).

**Figure 5:**
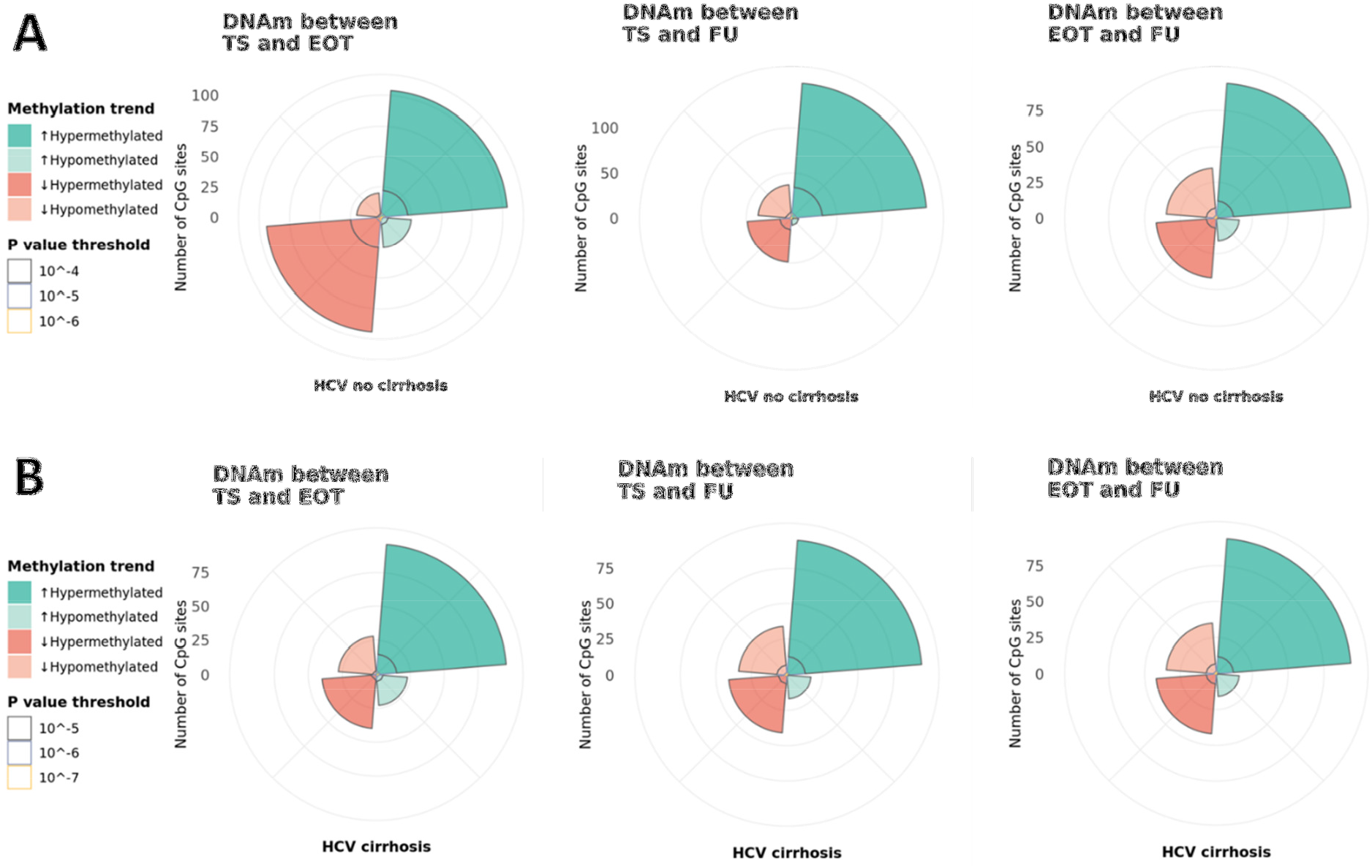
Global methylation trends in HCV patients without cirrhosis (cohort A) (A) and HCV patients with cirrhosis (cohorts B and C) (B). Color is indicating whether methylation increased (green) or decreased (red) and the overall methylation status of affected CpGs. Different sampling time points (TS = treatment start, EOT = end of treatment, FU= follow-up) were compared to show short-term and long-term effects.

**Figure 6:**
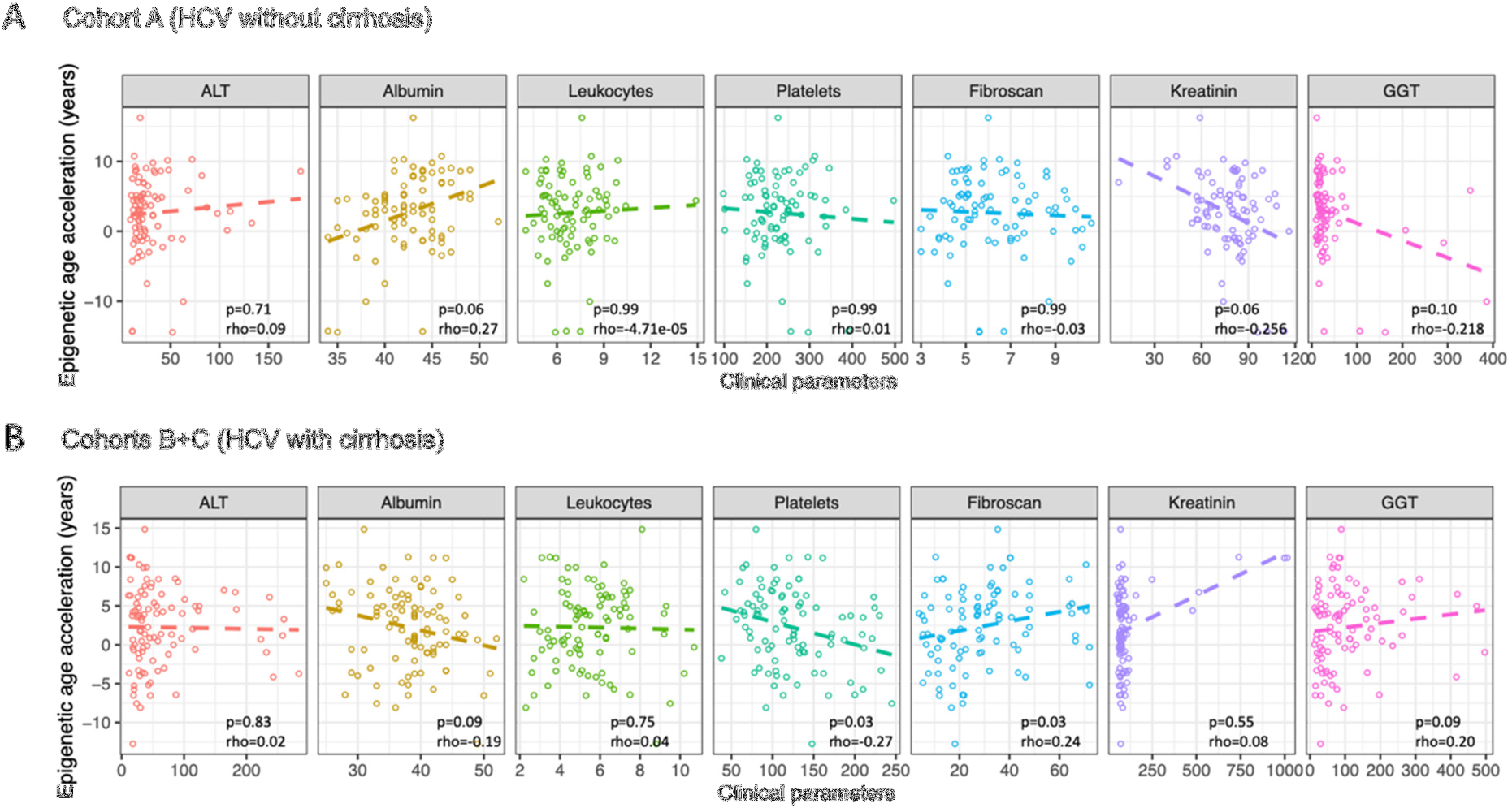
Scatter plots showing the correlation between clinical phenotype and epigenetic age acceleration in chronic HCV patients without cirrhosis (A) and with cirrhosis (B). P-values for correlations were adjusted using false discovery rate.

## Discussion

The results of our study suggest that chronic HCV infection leads to a general acceleration of epigenetic aging predicted by Horvath’s clock and that HCV elimination by DAA therapy can partially slow, halt, or reverse biological aging. The exact mechanism leading to this age acceleration and its reversal remains elusive. It has already been suggested that chronic infections such as HIV and HBV are associated with older or accelerated biological age [26] [27]. Thus, viral factors themselves may induce epigenetic modifications [28]. However, a recently published study did not observe this effect in monoinfected HCV patients. In this study, EAA was associated with advanced fibrosis and HIV coinfection in HCV patients. Of note, the HCV patients in the study by Gindin et al. were predominantly African American and were compared with the publicly available dataset consisting of Caucasian and Hispanic individuals [29]. Genetic background may potentially influence the results as African Americans seem to have a lower extrinsic epigenetic aging rate than Caucasians and Hispanics [30]. In addition, longitudinal data were not available and there was no information on ALT levels as marker for liver inflammation. Chronic inflammatory processes play a central role in the aging process, which is referred to as “inflammaging”, because inflammation promotes, among other things, the formation of reactive oxygen species that can cause DNA damage and thus also contribute to epigenetic changes [31].

Eliminating inflammation or the trigger of inflammation, such as with antiviral therapy, may possibly lead to slowing, halting or even reversing biological aging. Recently, it was shown that HIV patients showed improvement in epigenetic age after 96 weeks of antiretroviral therapy [32]. Gindin et al. showed that one-year antiviral treatment of chronic hepatitis B was associated with a modest reduction in age acceleration [29].

Our data also suggest that the process of EAA can in principle be halted or reversed, as after DAA therapy and HCV elimination, patients showed a significant increase in overall DNAm and a decrease in EAA predicted by the Horvath clock. However, this process appears to take time and is not immediately evident with the end of treatment. Also, not all patients show a decline to values of healthy controls. This is consistent with previous studies showing that HCV elimination does not always lead to complete clinical resolution [33] and also immunological imprints of HCV infection remain after SVR [8]. This may be particularly important in patients with advanced fibrosis or cirrhosis who have a residual risk of developing HCC after SVR [34] [35]. It has been demonstrated that HCV induced epigenetic changes in hepatocytes were associated with HCC and this persisted after SVR [13]. In addition, it has been shown that an accelerated age correlates with a higher risk of cancer associated mortality and overall mortality [36]. Consistent with this concept, in our study, patients who developed HCC after SVR had the highest EAA and showed no significant age deceleration after DAA treatment. This is important data not only to understand disease pathogenesis but there also remains an unmet need for high-quality markers for risk stratification of HCC development after SVR in former HCV patients. Early detection of HCC can lead to a decreased overall mortality. Our results suggest that age and deceleration offer new insights and could be a tool to improve conventional risk scores such as GALAD that comprise chronological age [37]. Instead of using chronological age, which is an imperfect surrogate measure of the aging process [15], biological age may ultimately better reflect the risk of developing HCC. Thus, DNA methylation analysis of peripheral blood cells could serve as biomarker in terms of a liquid biopsy to improve the management of patients with chronic hepatitis C or other patients with chronic inflammatory conditions. For the development of a biomarker, it is important to consider that aging is accompanied by a change in blood cell type composition, e.g., the proportion of naïve or senescent cytotoxic T cells changes with age, which could create a bias. The age calculated by the Horvath clock seems to be largely unaffected by these changes [15], which is consistent with our data as the changes in DNA methylation were not significantly associated with altered cell type composition.

Our study certainly has strengths and limitations. The strengths lie in a very well characterized and matched clinical cohort of 54 patients in total. The cohort of patients who developed HCC is very unique as we could analyze patient samples before the development of HCC. As the number of HCC patients was limited, larger studies are needed to evaluate whether the dynamics of age acceleration can support clinical decision making in risk stratification for developing HCC in chronic HCV patients.

In conclusion, our study contributes to the understanding of the biological effects of HCV elimination after DAA therapy and offers new conceptual ideas for the use of DNAm in peripheral blood cells as a biomarker that supports the current effort for more individualized infectious disease medicine.

## Data Availability

All data produced in the present study are available upon reasonable request to the authors

## Abbreviations

ALT: Alanine amino transferase
AST: Aspartate aminotransferase
CpG: 5’-C-phosphate-G-3’
DAA: Direct-acting antiviral
DMSO: Dimethyl sulfoxide
DNA: Deoxyribonucleic acid
DNAm: Deoxyribonucleic acid methylation
EAA: Epigenetic age acceleration
FCS: Fetal calf serum
GALAD: Gender, age, AFP-L3, AFP and Des-carboxy-prothrombin (DCP)
GGT: Gamma-glutamyl transferase
HBV: Hepatitis B virus
HCC: Hepatocellular carcinoma
HCV: Hepatitis C virus
HIV: Human immunodeficiency virus
INR: International normalized ratio
PBMC: Peripheral blood mononuclear cell
PS: Propensity score
RNA: Ribonucleic acid
RPMI: Roswell Park Memorial Institute medium
SNP: Single-nucleotide polymorphism
SOP: Standard Operation Procedure
SVR: Sustained virological response

## Acknowledgments

We thank Helena Lickei and Hagen Schmaus for their assistance with blood sample processing. We thank the study nurses (Neslihan Devici, Carola Mix, Janet Cornberg, Jennifer Witt, Julia Schneider) and the physicians (Katja Deterding, Christopher Dietz, Kerstin Port, Tammo Tergast) of the Hepatitis Outpatient Clinic of the Department of Gastroenterology, Hepatology and Endocrinology of Hannover Medical School for the care of the patients in the patient registry. We thank all patients for participating in our research study and for donating blood. MU was supported by the Hannover Biomedical Research School (HBRS) and the Center for Infection Biology (ZIB).

## Supplementary information

## Supplementary Tables

**[Supplementary Table 1:**
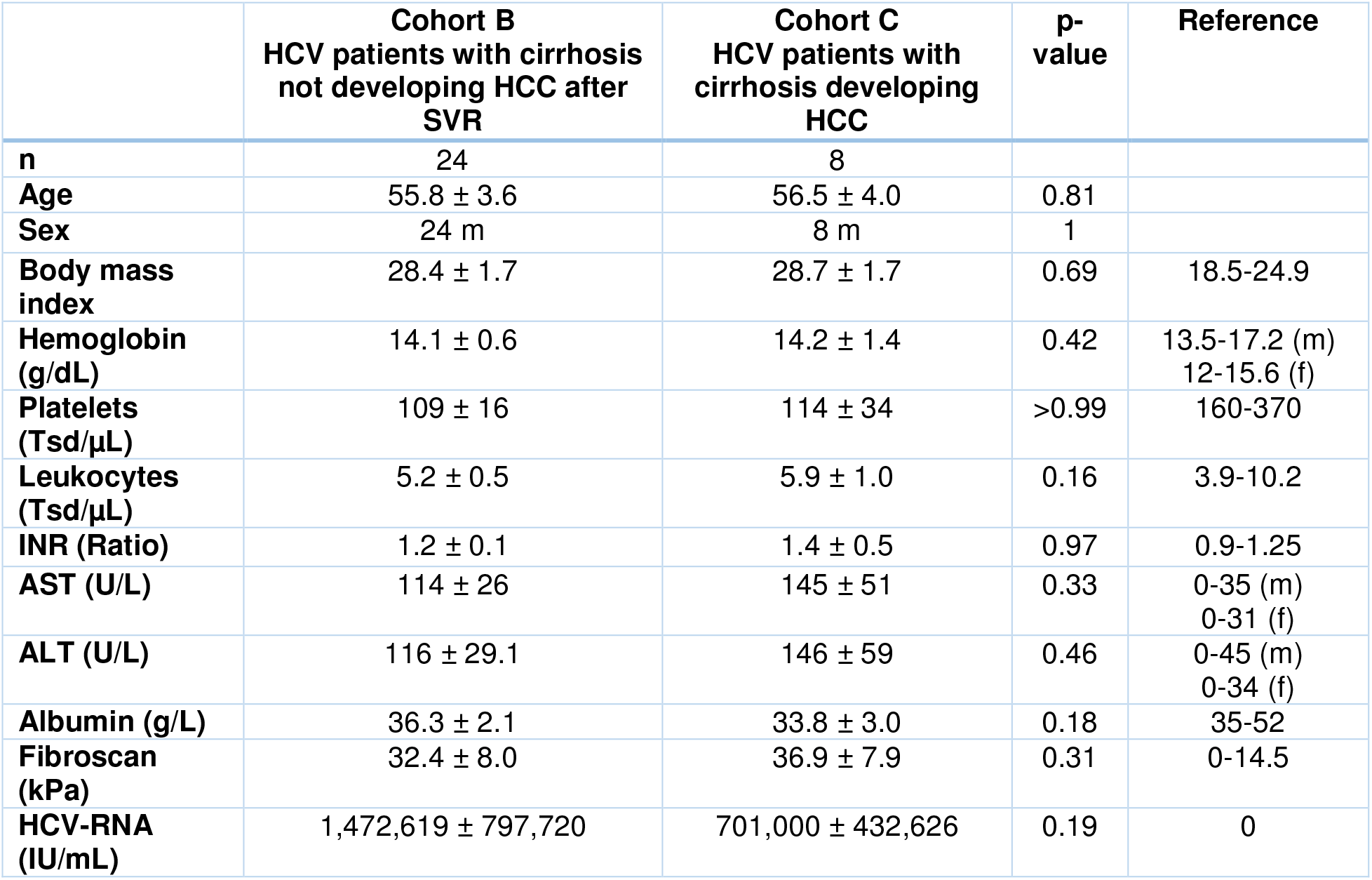
Baseline characteristics and comparison between PS-matched cohorts B and C.

|  | **Cohort B**<br>HCV patients with cirrhosis<br>not developing HCC after<br>SVR | **Cohort C**<br>HCV patients with<br>cirrhosis developing<br>HCC | **p-**<br>**value** | **Reference** |
| --- | --- | --- | --- | --- |
| **n** | 24 | 8 |  |  |
| **Age** | 55.8 ± 3.6 | 56.5 ± 4.0 | 0.81 |  |
| **Sex** | 24 m | 8 m | 1 |  |
| **Body mass**<br>**index** | 28.4 ± 1.7 | 28.7 ± 1.7 | 0.69 | 18.5-24.9 |
| **Hemoglobin**<br>**(g/dL)** | 14.1 ± 0.6 | 14.2 ± 1.4 | 0.42 | 13.5-17.2 (m)<br>12-15.6 (f) |
| **Platelets**<br>**(Tsd/μL)** | 109 ± 16 | 114 ± 34 | >0.99 | 160-370 |
| **Leukocytes**<br>**(Tsd/μL)** | 5.2 ± 0.5 | 5.9 ± 1.0 | 0.16 | 3.9-10.2 |
| **INR (Ratio)** | 1.2 ± 0.1 | 1.4 ± 0.5 | 0.97 | 0.9-1.25 |
| **AST (U/L)** | 114 ± 26 | 145 ± 51 | 0.33 | 0-35 (m)<br>0-31 (f) |
| **ALT (U/L)** | 116 ± 29.1 | 146 ± 59 | 0.46 | 0-45 (m)<br>0-34 (f) |
| **Albumin (g/L)** | 36.3 ± 2.1 | 33.8 ± 3.0 | 0.18 | 35-52 |
| **Fibroscan**<br>**(kPa)** | 32.4 ± 8.0 | 36.9 ± 7.9 | 0.31 | 0-14.5 |
| **HCV-RNA**<br>**(IU/mL)** | 1,472,619 ± 797,720 | 701,000 ± 432,626 | 0.19 | 0 |

## Supplementary Figures

**[Supplementary Figure 1:**
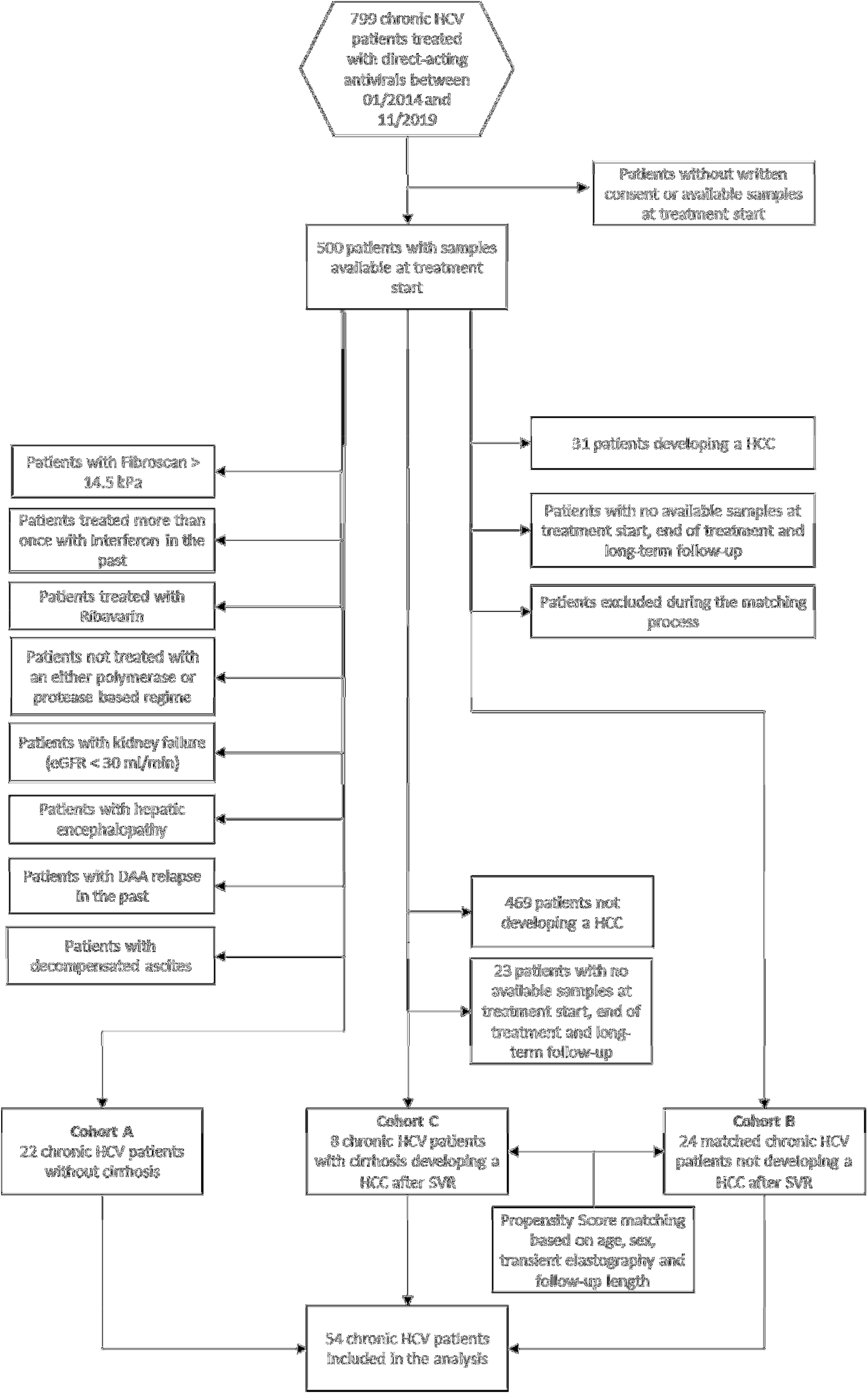
Inclusion and exclusion criteria for analyzed HCV patients

**[Supplementary Figure 2:**
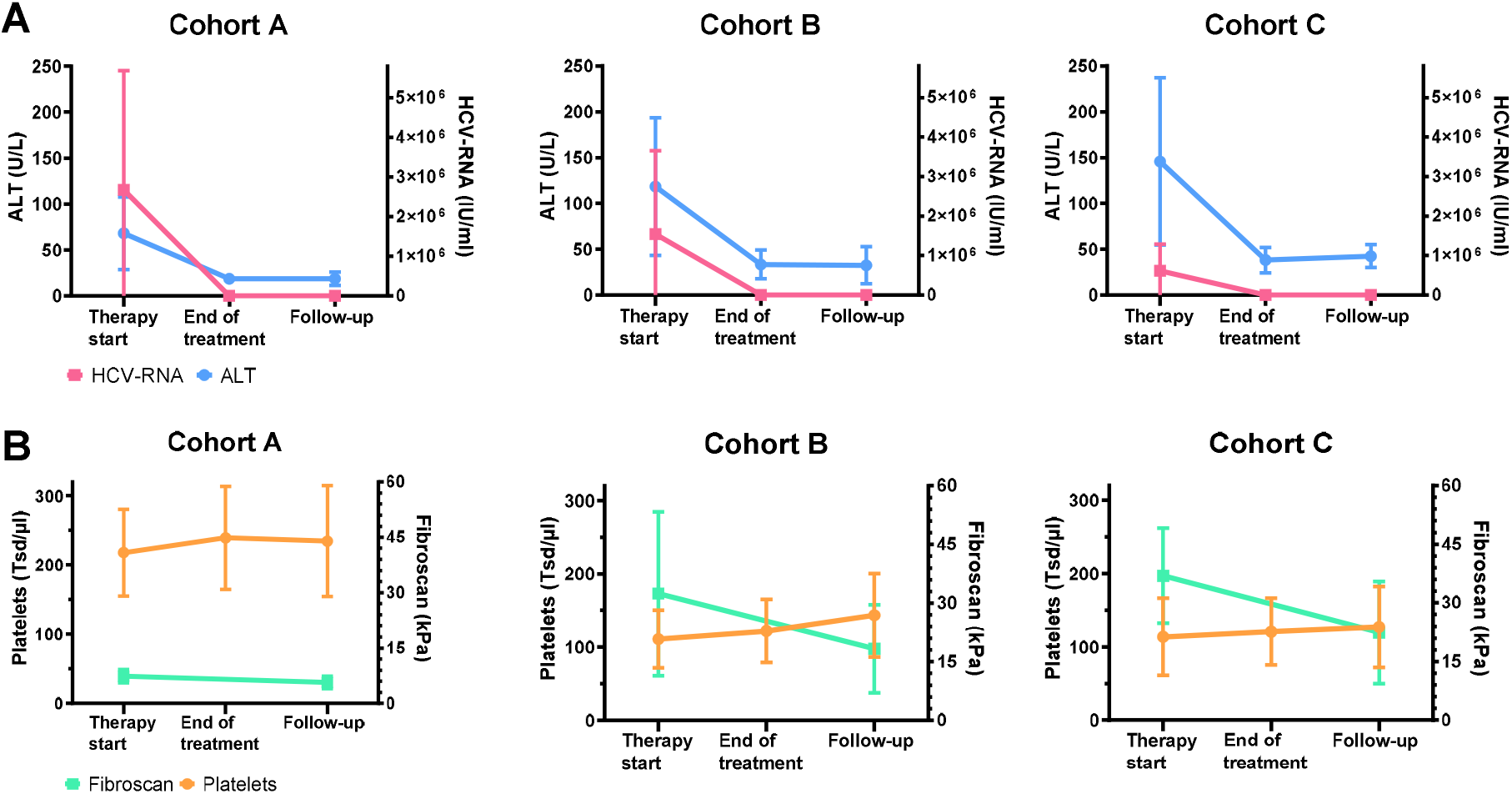
HCV-RNA, ALT (A), transient elastography (FibroScan) and platelets levels (B) before, after treatment and at follow-up in the three cohorts (cohort A = no cirrhosis [n=22], cohort B = cirrhosis without HCC [n=24], cohort C = cirrhosis with HCC [n=8]). All errorbars indicate standard deviations.

**[Supplementary Figure 3:**
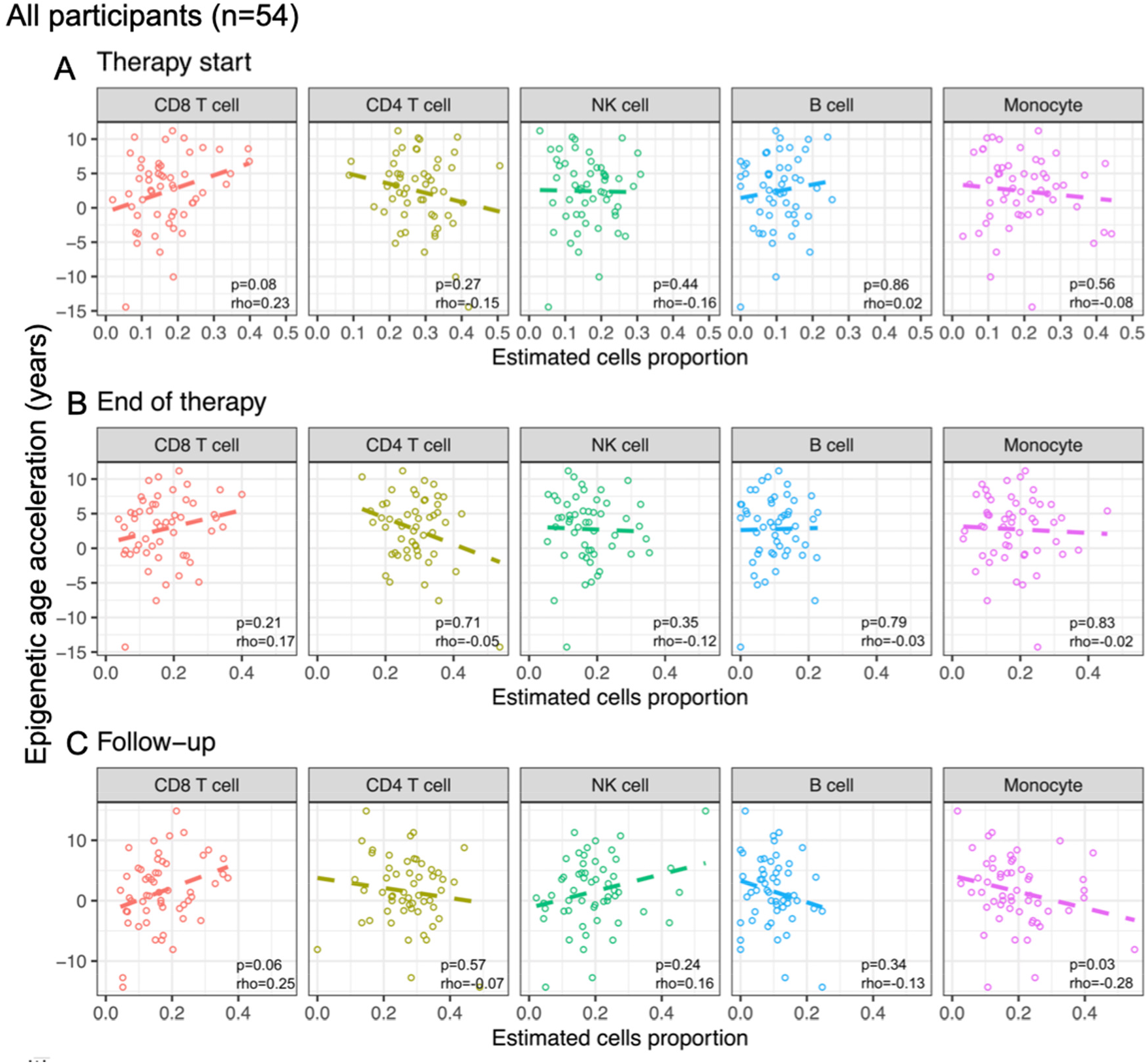
Correlation between cell type estimates and epigenetic age acceleration in chronic HCV patients. Correlations are shown at “therapy start” (A), “end of treatment” (B) and “follow-up” (C).

**[Supplementary Figure 4:**
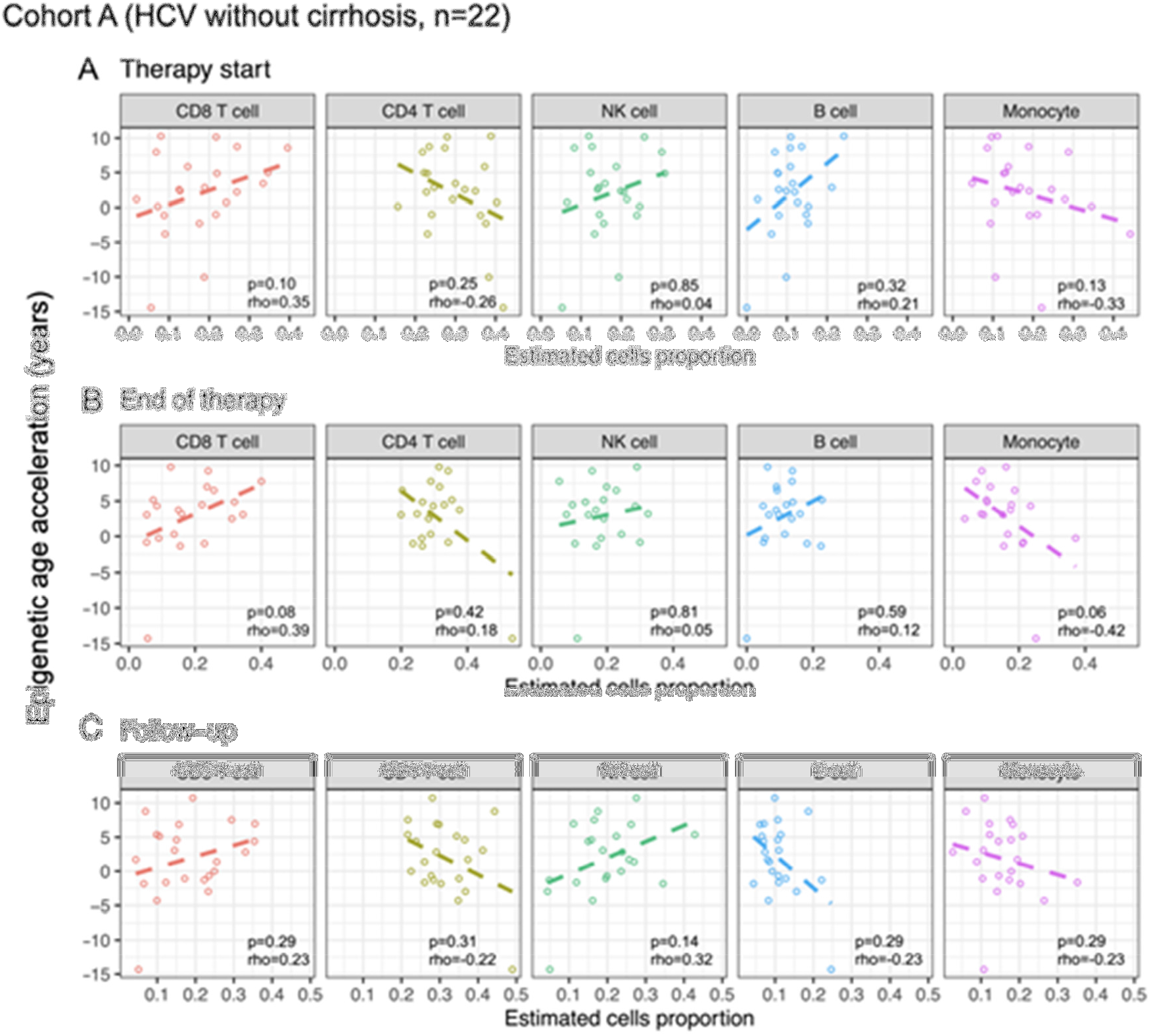
Correlation between cell type estimates and age acceleration in chronic HCV patients without cirrhosis (cohort A). Correlations are shown at “therapy start” (A), “end of treatment” (B) and “follow-up” (C).

**[Supplementary Figure 5:**
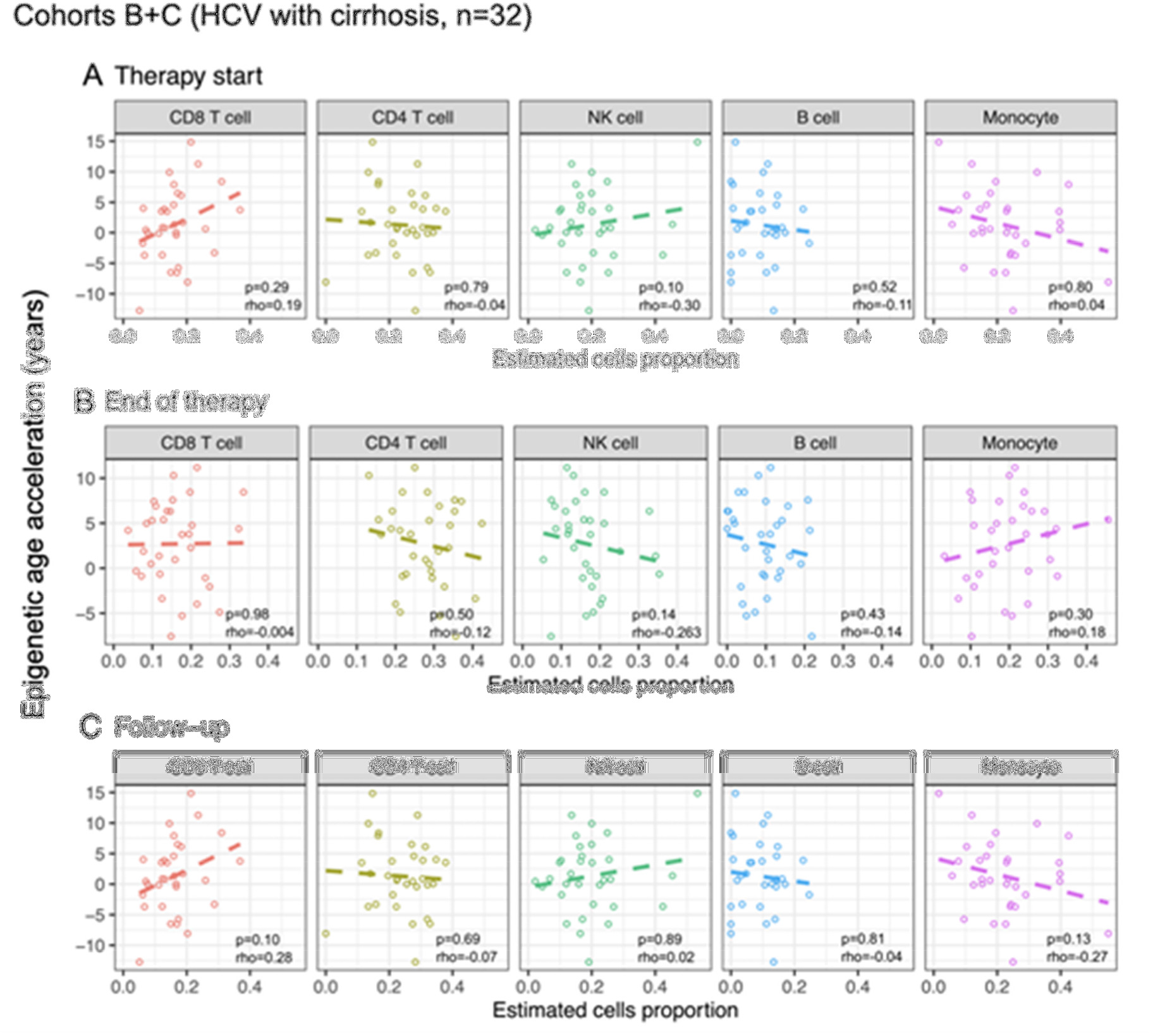
Correlation between cell type estimates and age acceleration in chronic HCV patients with cirrhosis (cohorts B and C). Correlations are shown at “therapy start” (A), “end of treatment” (B) and “follow-up” (C).

## References

[1] World Health Organization. Hepatitis C: Key facts 2021.

[2] Westbrook R H, Dusheiko G. Natural history of hepatitis C. Journal of hepatology 2014; 61:S58–S68.

[3] Cacoub P, Saadoun D. Extrahepatic Manifestations of Chronic HCV Infection. The New England journal of medicine 2021; 384:1038–1052.

[4] van der Meer, Adriaan J, Berenguer M. Reversion of disease manifestations after HCV eradication. Journal of hepatology 2016; 65:S95–S108.

[5] Negro F. Residual risk of liver disease after hepatitis C virus eradication. J Hepatol 2021; 74:952–963.

[6] Ohlendorf V, Schäfer A, Christensen S, Heyne R, Naumann U, Link R et al. Only partial improvement in health-related quality of life after treatment of chronic hepatitis C virus infection with direct acting antivirals in a real-world setting–results from the German Hepatitis C-Registry (DHC-R). Journal of Viral Hepatitis 2021.

[7] Kondili L A, Monti M, Quaranta MG, Gragnani L, Panetta V, Brancaccio G et al. A prospective study of DAA Effectiveness and Relapse Risk in HCV Cryoglobulinemic Vasculitis by the Italian PITER Cohort. Hepatology (Baltimore, Md.) 2021.

[8] Hensel N, Gu Z, Sagar, Wieland D, Jechow K, Kemming J et al. Memory-like HCV-specific CD8+ T cells retain a molecular scar after cure of chronic HCV infection. Nat Immunol 2021; 22:229.

[9] Aregay A, Owusu Sekyere S, Deterding K, Port K, Dietz J, Berkowski C et al. Elimination of hepatitis C virus has limited impact on the functional and mitochondrial impairment of HCV-specific CD8+ T cell responses. Journal of Hepatology 2019; 71:889–899.

[10] Strunz B, Hengst J, Deterding K, Manns MP, Cornberg M, Ljunggren H et al. Chronic hepatitis C virus infection irreversibly impacts human natural killer cell repertoire diversity. Nature communications 2018; 9:2275.

[11] Yates K B, Tonnerre P, Martin GE, Gerdemann U, Al Abosy R, Comstock DE et al. Epigenetic scars of CD8+ T cell exhaustion persist after cure of chronic infection in humans. Nature immunology 2021; 22:1020–1029.

[12] Hlady R A, Zhao X, El Khoury LY, Luna A, Pham K, Wu Q et al. Interferon drives HCV scarring of the epigenome and creates targetable vulnerabilities following viral clearance. Hepatology (Baltimore, Md.) 2021; 00:1.

[13] Hamdane N, Jühling F, Crouchet E, El Saghire H, Thumann C, Oudot MA et al. HCV-Induced Epigenetic Changes Associated With Liver Cancer Risk Persist After Sustained Virologic Response. Gastroenterology (New York, N.Y. 1943) 2019; 156:2313–2329.e7.

[14] Morales-Nebreda L, McLafferty FS, Singer BD. DNA methylation as a transcriptional regulator of the immune system. Translational research : the journal of laboratory and clinical medicine 2019; 204:1–18.

[15] Horvath S, Raj K. DNA methylation-based biomarkers and the epigenetic clock theory of ageing. Nature Reviews Genetics 2018; 19:371–384.

[16] Xu C, Bonder MJ, Söderhäll C, Bustamante M, Baïz N, Gehring U et al. The emerging landscape of dynamic DNA methylation in early childhood. BMC genomics 2017; 18:25.

[17] Horvath H, Horvath S. DNA methylation age of human tissues and cell types DNA methylation age of human tissues and cell types. Genome Biology 2013; 14.

[18] Ho D E, Imai K, King G, Stuart EA. MatchIt: Nonparametric Preprocessing for Parametric Causal Inference. Journal of statistical software 2011; 42.

[19] Randolph J J, Austin KF, Manuel K, Balloun JL, Randolph JJ;, Falbe K; et al. A Step-by-Step Guide to Propensity Score Matching in R. Practical Assessment, Research, and Evaluation 2014; 19.

[20] Aryee M J, Jaffe AE, Corrada-Bravo H, Ladd-Acosta C, Feinberg AP, Hansen KD et al. Minfi: a flexible and comprehensive Bioconductor package for the analysis of Infinium DNA methylation microarrays. Bioinformatics 2014; 30:1363–1369.

[21] Pidsley R, Zotenko E, Peters TJ, Lawrence MG, Risbridger GP, Molloy P et al. Critical evaluation of the Illumina MethylationEPIC BeadChip microarray for whole-genome DNA methylation profiling. Genome Biol 2016; 17.

[22] Illumina Inc. BeadArray Controls Reporter Software Guide 2015.

[23] Pidsley R, Y Wong CC, Volta M, Lunnon K, Mill J, Schalkwyk LC. A data-driven approach to preprocessing Illumina 450K methylation array data. BMC genomics 2013; 14:293.

[24] Hannum G, Guinney J, Zhao L, Zhang L, Hughes G, Sadda S et al. Genome-wide Methylation Profiles Reveal Quantitative Views of Human Aging Rates. Molecular cell 2013; 49:359–367.

[25] Heyn H, Li N, Ferreira HJ, Moran S, Pisano DG, Gomez A et al. Distinct DNA methylomes of newborns and centenarians. Proceedings of the National Academy of Sciences - PNAS 2012; 109:10522–10527.

[26] Gross A, Jaeger P, Kreisberg J, Licon K, Jepsen K, Khosroheidari M et al. Methylome-wide Analysis of Chronic HIV Infection Reveals Five-Year Increase in Biological Age and Epigenetic Targeting of HLA. Molecular cell 2016; 62:157–168.

[27] Horvath S, Levine AJ. HIV-1 Infection Accelerates Age According to the Epigenetic Clock. The Journal of infectious diseases 2015; 212:1563–1573.

[28] Tachiwana H, Shimura M, Nakai-Murakami C, Tokunaga K, Takizawa Y, Sata T et al. HIV-1 Vpr Induces DNA Double-Strand Breaks. Cancer Res 2006; 66:627.

[29] Gindin Y, Gaggar A, Lok AS, Janssen HLA, Ferrari C, Subramanian GM et al. DNA Methylation and Immune Cell Markers Demonstrate Evidence of Accelerated Aging in Patients with Chronic Hepatitis B Virus or Hepatitis C Virus, with or without Human Immunodeficienct Virus Co-infection. Clinical infectious diseases 2021; 73:e184–e190.

[30] Horvath S, Gurven M, Levine ME, Trumble BC, Kaplan H, Allayee H et al. An epigenetic clock analysis of race/ethnicity, sex, and coronary heart disease. Genome Biol 2016; 17.

[31] Ohnishi S, Ma N, Thanan R, Pinlaor S, Hammam O, Murata M et al. DNA Damage in Inflammation-Related Carcinogenesis and Cancer Stem Cells. Oxidative medicine and cellular longevity 2013; 2013:387014–9.

[32] Esteban-Cantos A, Rodríguez-Centeno J, Saiz-Medrano G, Bsc M, De Miguel R, Bernardino JI et al. Articles Epigenetic age acceleration changes 2 years after antiretroviral therapy initiation in adults with HIV: a substudy of the NEAT001/ANRS143 randomised trial 2019.

[33] Loo N, Hanysak B, Mann J, Ramirez R, Kim J, Mitchell R et al. Real-world observational experience with direct-acting antivirals for hepatitis C: baseline resistance, efficacy, and need for long-term surveillance. Medicine (Baltimore) 2019; 98:e16254.

[34] Kanwal F, Kramer JR, Asch SM, Cao Y, Li L, El-Serag HB. Long-Term Risk of Hepatocellular Carcinoma in HCV Patients Treated With Direct Acting Antiviral Agents. Hepatology 2020; 71.

[35] Waziry R, Hajarizadeh B, Grebely J, Amin J, Law M, Danta M et al. Hepatocellular carcinoma risk following direct-acting antiviral HCV therapy: A systematic review, meta-analyses, and meta-regression. J Hepatol 2017; 67:1204–1212.

[36] Perna L, Zhang Y, Mons U, Holleczek B, Saum K, Brenner H. Epigenetic age acceleration predicts cancer, cardiovascular, and all-cause mortality in a German case cohort. Clinical epigenetics 2016; 8:64.

[37] Best J, Bilgi H, Heider D, Schotten C, Manka P, Bedreli S et al. The GALAD scoring algorithm based on AFP, AFP-L3, and DCP significantly improves detection of BCLC early stage hepatocellular carcinoma. Z Gastroenterol 2016; 54:1296.

